# Post-Surgical Scar Management and Rehabilitation in Burn Patients: Insights from Gaza’s challenging context – A Retrospective Descriptive Study

**DOI:** 10.64898/2026.03.10.26348110

**Authors:** Abed El Hamid Qaradaya, Julie Van Hulse, Jomaa Younis, Feras Swairjo, Hala Al Far, Noura Al Zaeem, Ebtihal Wally, Mohamad Abu Hashem, Rami Al Shamali, Mabade Al Farra, Natalie Thurtle, Mohammed Abu Mughiaseeb, Mohamad Al Ghazali, Jihane Ben Farhat

## Abstract

**Background:** In the conflict-ridden Gaza Strip, burn injuries pose significant challenges for affected individuals. Often stemming from accidents involving generators and substandard gas cylinders, these burns can have profound physical and psychological impacts. Access to comprehensive medical care is limited in this context, making specialized burn treatment centers critical lifelines for those in need. This study seeks to explore the outcomes of rehabilitation and physiotherapy for post-surgical skin grafts in specialized burn treatment centers in Gaza, recognizing the crucial role such interventions play in improving the quality of life for burn survivors in this challenging environment.

**Methods:** We carried out a retrospective descriptive study of patients with post-surgical skin graft (SSG) scars subsequent to burns injury, and enrolled in the MSF post-op outpatient clinics in the Gaza Strip (Gaza City, Beit Lahia, and Khan Younis), between January 2018 and December 2020. Our analysis used five outcome measures to assess the effects of physiotherapy and post-surgical treatment: the reducibility score, which indicates the extent to which scars can be reduced in size or prominence; the Functional Activity for Burn (FAB) score, which evaluates the functional capabilities of patients following burn injury; the Vancouver Scar Scale (VSS), which assesses various aspects of scar appearance such as pigmentation, vascularity, pliability, and height; the Visual Analgesic Scale (VAS), which quantifies pain levels experienced by patients; and the itching score, which measures the severity of itching associated with scar formation

**Results:** A total of 177 patient records were examined, revealing that the majority of burn victims in Gaza were children under the age of 18 (n=136, 76.8%), with scalding from liquid burns being the primary cause (n=119, 67.6%). The outcomes of the physiotherapy program varied depending on the type of pressure therapy and insert material utilized. However, significant improvements were observed in various outcome measures following enrollment in the program. These improvements included reductions in pain scores (mean initial score: 5.3, SD: 2.5; mean final score: 1.4, SD: 1.8), itching (mean initial score: 3.7, SD: 2.7; mean final score: 2.7, SD: 2.2), scar pigmentation, vascularity, pliability, and height (mean initial VSS: 7.1, SD: 1.8; mean final VSS: 5.7, SD: 1.7). Moreover, treatment correlated with enhancements in overall function (mean initial score: 25.6, SD: 7.1; mean final score: 34.6, SD: 2.3) and a reduction in skin contracture (mean initial score: 2.3, SD: 1.4; mean final score: 0.8, SD: 0.9).

**Conclusions:** Our findings underscore the important role of rehabilitation and physiotherapy in optimizing outcomes for post-surgical skin graft patients in conflict-affected regions like the Gaza Strip. Despite the challenging environment, MSF’s clinic in Gaza has demonstrated the feasibility and effectiveness of delivering comprehensive care to burn survivors. Moving forward, further research is needed to refine and validate best practices in rehabilitation interventions tailored to the specific needs of patients in conflict zones, ensuring continued progress in enhancing the quality of care and quality of life for those affected by burn injuries.

## BACKGROUND

The Gaza Strip, nestled amidst ongoing socio-political strife, stands as an emblem of the enduring repercussions of conflict. Characterized by its constrained geographical expanse and a staggering population density of 5,046 inhabitants per km2, this territory grapples with multifaceted challenges exacerbated by socio-political strife. The blockade imposed by Israel has further exacerbated the plight of its inhabitants, with over a million people – nearly half the population – pushed below the poverty line in just over a decade (6,7). Within this backdrop, injuries, both domestic and conflict-related, impose a heavy toll, underscoring the critical need for robust and adaptive care strategies. In Gaza, the aftermath of burns transcends mere physical trauma, often culminating in incapacitating hypertrophic scars. Yet, scar management techniques, especially in the critical post-surgical phase, persist as neglected territories, particularly in pediatric cases (5). Scar management techniques, especially post-surgical, remain underexplored, particularly in pediatric cases and in such contexts. Amidst socio-political challenges, Médecins Sans Frontières (MSF) has been providing vital care, including specialized physiotherapy and rehabilitation services.

Hypertrophic scars result from an overabundance of fibroblast-produced extracellular matrix proteins, notably collagen, persisting over extended durations. These scars pose significant risks, including skin contractures and impairments in both range of motion and functional capacity. Skin graft procedures similarly predispose individuals to hypertrophic scarring, exacerbating these enduring complications. Numerous therapeutic approaches have been devised to address post-burn scarring, each subjected to rigorous evaluation through research endeavors. A systematic review of diverse studies indicates that employing pressure garments for a minimum of 23 hours daily over 6–12 months, with pressures ranging from 20 to 40 mmHg, can effectively enhance scar mobility in joint regions, forestall contractures, and mitigate the risk of scar contraction (2). Additionally, the utilization of silicone sheets and gels is advocated for both the prevention and treatment of hypertrophic scars and keloids. Research underscores the significant benefits of topical silicone sheets, manifesting in notable improvements in the color, thickness, and suppleness of hypertrophic scars and keloids (3).

Few research has delved into the efficacy of scar management techniques, especially concerning post-surgical scars, particularly among pediatric populations. Treating burns in children presents unique challenges owing to their developmental stage and rapid growth rate. Scar treatment in this demographic necessitates a comprehensive rehabilitation regimen, encompassing diverse modalities such as scar tissue manipulation, joint mobilization, functional rehabilitation to facilitate daily activities, and utilization of various compression tools and materials. This holistic approach is pivotal in curbing scar progression over the protracted maturation period, which can extend up to two years.

Médecins Sans Frontières (MSF) started working in the Occupied Palestinian Territories in 1989, to treat victims of the Israeli-Palestinian conflict. In 2000, MSF began activity in Gaza with a psychological project and in 2007, then launched a medical and rehabilitation project to treat victims of trauma and burns. For the last 14 years the project has treated multiple forms of trauma including injuries linked to violence, domestic burns injuries, and hand injuries. A physiotherapy and rehabilitation department forms part of the programme, providing rehabilitation through a multidisciplinary team. In 2020, 11,330 physiotherapy sessions were provided to burns patients, with an average of 14 sessions per patient over the course of treatment. In the same year, 89 burn patients received skin graft surgery from the MSF clinic. Most patients in this project are children who have experienced domestic burns accidents. MSF has a specialized physiotherapy and rehabilitation protocol for treating burns patients; the protocol focuses on providing scar tissue massage (using gradual divergent, multi-axis two-handed massage, detaching and folding the skin, and skin release and torsion), skin stretching techniques, functional rehabilitation and mobility exercises, scar tissue hydration using cold creams, and compression therapy involving pressure garments, cohesive bandages, and tubular elastic bandage. Finally, insert materials are also used such as silicon Sheets, soft putty elastomer, and polyethylene foam (Plastazote).

We aimed to describe the patients who received rehabilitation/physiotherapy care for post-surgical skin grafts. We also aim to describe the use of materials like silicon and soft putty for scar management and outline the characteristics and outcomes of these patients.

## METHODS

### Study site

The study took place at Burn Center which had been established in 2007 and is located in Nasser Medical Complex in Gaza Strip. It serves more than 1.1 million inhabitants and covers all Gaza governorates except south governorates. It has 12 beds distributed in 6 rooms in addition to a dressing and hydrotherapy room. Furthermore, three ICU beds and one operating room are available in service. Care is provided by plastic surgeons, nurses and physiotherapists, antimicrobial stewardship doctors, as well as psychological support from psychologists.

### Design

This study employed a retrospective descriptive design, involving patients who underwent skin graft surgery as part of MSF’s physiotherapy and rehabilitation program in the Gaza Strip from January 1st, 2018, to December 31st, 2020. Eligible patients had post-surgical skin graft (SSG) scars resulting from burns during the study period and were discharged from clinic. To assess improvement after physiotherapy treatment, five outcome measures were utilized. Firstly, the Reducibility score measured the extent of contractures resulting from skin pulling together after second- or third-degree burns, ranging from 0 (better) to 4 (worse). Secondly, the Functional Activity for Burn (FAB) score evaluated patients’ functional abilities, ranging from 7 (worse) to 35 (better). Thirdly, the Vancouver Scar Scale (VSS) assessed scar improvement in terms of pigmentation, vascularity, pliability, and height, with scores ranging from 0 (better) to 13 (worse). Fourthly, pain scores were recorded using the Visual Analogue Scale (VAS), ranging from 0 (no pain) to 10 (most unbearable pain), comparing pain levels before and after treatment. Finally, itching scores evaluated scar itchiness before and after treatment, ranging from 0 (no itching) to 10 (very itchy).

### Data collection

Data were accessed for research purposes between August and October 2021. All data were extracted from routine clinical records and anonymized prior to analysis. The authors did not have access to information that could identify individual participants during or after data collection. Data were entered into RedCap (Vanderbuilt University ©). The demographic information gathered for each patient encompassed age, sex, employment, and educational status, as well as any post-burn related comorbidities. Additionally, details regarding injury characteristics, including cause, location, type, pain levels, reducibility scores, scar assessments, and surgical interventions such as operations and procedures, along with their timing, were documented. Furthermore, information on the specific physiotherapy and rehabilitation modalities employed for each patient was recorded, encompassing the treatment scheme, utilized materials (such as silicone sheets, soft putty elastomer, and polyethylene foam), pain management strategies, itching management, treatment frequency and timing, as well as injury and episode outcomes.

### Statistical analysis

We analyzed the data descriptively, providing details on the median and inter-quartile range (IQR) for numeric variables and proportions for categorical variables. The total number of non-missing values served as the denominator for calculations. Additionally, differences between outcomes before and after treatment were calculated, including mean difference and standard deviation. Data was analyzed using Stata V.15 software (Stata Corporation, Texas, USA).

## RESULTS

### Demographics and Subject Characteristics

A total of 240 patients met the inclusion criteria, having undergone skin graft surgery between January 1st, 2018, and December 31st, 2020, and completed their physiotherapy treatment course (Table 1). However, 63 patients had incomplete data and were therefore excluded from the analysis. Consequently, the analysis included 177 patients. The study population was young, with a median age of 5 years (IQR 2-13), and the majority were under 18 years old (n=136; 76.8%). Males accounted for 56.5% of the population (n=100). The prevalence of post-burn related comorbidities was low, with only one patient having undergone a fasciotomy. Over 70% of the study population sustained minor burns (children: n=95, 73.1%; adults: n=28, 70%), predominantly caused by scalds (n=119, 67.2%) (Supplementary material). Most injuries affected the upper limbs (n=106, 56.9%), with corresponding graft sites primarily located on the upper limbs (n=91, 52.4%) (Table 2). The thigh area was the most common donor skin site (n=167, 49.3%) (Table 2).

**Table 1:** Characteristics of study population.

| Characteristics |  | N=177 |  |
| --- | --- | --- | --- |
| Age, median [IQR], years |  | 5 | [2-13] |
| Age groups in years, n (%) |  |  |  |
|  | <1 | 29 | (16.4) |
|  | 1-5 | 63 | (35.6) |
|  | 6-10 | 31 | (17.5) |
|  | 11-19 | 15 | (8.5) |
|  | ≥20 | 39 | (22.0) |
| Child vs Adult, n (%) |  |  |  |
|  | Child 0-18 y.o. | 136 | (76.8) |
|  | Adult 19+ | 41 | (23.2) |
| Gender, n (%) |  |  |  |
|  | Female | 77 | (43.5) |
|  | Male | 100 | (56.5) |
| Comorbidities, n (%) |  |  |  |
|  | Fasciotomy | 1 | (0.6) |
|  | None | 176 | (99.4) |

**Table 2:** Location of burns and sites of graft.

|  | Location of burns<br>n (%) | Sites of grafts<br>n (%) | Donor site | n (%) |
| --- | --- | --- | --- | --- |
| Left Upper limb | 50 (25.3) | 46 (26.0) | Left thigh | 79 (44.6) |
| Right upper limb | 56 (31.6) | 45 (25.4) | Right thigh | 88 (49.7) |
| Left hand | 24 (13.6) | 17 (9.6) | Left leg | - |
| Right hand | 28 (15.8) | 25 (14.1) | Right leg | 3 (1.7) |
| Left lower limb | 42 (23.7) | 35 (19.8) | Trunk | 6 (3.4) |
| Right lower limb | 42 (23.7) | 35 (19.8) | Other | 16 (9.0) |
| Anterior trunk | 33 (18.6) | 23 (13.0) |  |  |
| Posterior trunk | 14 (7.9) | 7 (4.0) |  |  |
| Face and head | 14 (7.9) | 6 (3.4) |  |  |

The mean delay between surgery and admission for physiotherapy was 13.2 days, while the average duration of physiotherapy treatment was 178.9 days, equivalent to approximately 5.9 months.

### Primary outcomes

Supplementary table shows physiotherapists predominantly relied on tube grip (77 patients, 44.5%) and pressure garments (61 patients, 35.3%) as primary methods for pressure therapy treatment. Among these, a majority of patients (40 patients, 82.4%) received pressure therapy alone, without any additional insert material (Supplementary Figure). Conversely, 146 patients underwent pressure therapy in conjunction with insert materials. Silicon sheets emerged as the most frequently utilized insert material (92 patients, 63%), followed by polyethylene foam plastizote® (45 patients, 30.8%), with only 9 patients receiving soft putty (6.1%) (Refer to Figure 3). On average, insert materials were utilized for a duration of 119.8 days (SD=82.5).

The majority of patients attended on at least 3 physiotherapy sessions per week (131, 74.0%) while fewer patients (39, 22%) attended 2 sessions per week. Most patients were discharged as cured (153, 86.4%), however 23 patients were discharged as defaulters and were lost to follow up (13.0%).

In total, 181 different types of pressure methods were used, with some patients receiving two methods. In general the clinic physiotherapists relied on use of tube grip (77, 43.5%) and pressure garments (61, 34.5%). Among those patients receiving only one pressure therapy, 35 patients had no insert materials used (21.2%). However, for 146 patients, an insert material was used under pressure therapy. The most commonly used type of insert material was silicon sheet (92, 63%) then polyethylene foam (Plastazote®; 45, 30.8%); for9 patients, soft putty elastomer was used (6.1%). Insert material was used for a mean of 119.8 days (SD, 82.5).

The findings in tables S4-S8 indicate that post-surgical skin graft physiotherapy led to improvements across various scar characteristics. The Vancouver Scar Scale (VSS) demonstrated a reduction in scar pigmentations, vascularity, and height, with enrolled patients showing a decrease from a mean initial VSS of 7.1 (SD, 1.8) to a mean final of 5.7 (SD, 1.7). Additionally, there was a notable decrease in pain levels, as reflected by the Visual Analogue Scale (VAS) scores, declining from a mean initial VAS of 5.3 (SD, 2.5) to a mean final score of 1.4 (SD, 1.8). Furthermore, patients experienced a reduction in scar itching, with the mean initial itching score decreasing from 3.7 (SD, 2.7) to a mean final score of 2.7 (SD, 2.2). Scar contracture and reducibility also improved, as evidenced by a decline in the Reducibility score from a mean initial score of 2.3 (SD, 1.4) to a mean final score of 0.8 (SD, 0.9). Moreover, there was a notable enhancement in functional abilities among enrolled patients, as measured by the Functional Activity for Burn (FAB) score, which increased from a mean initial score of 25.6 (SD, 7.1) to a mean final score of 34.6 (SD, 2.3).

### Secondary outcomes

The provided data in table 3 illustrate the outcomes associated with different insert materials used in pressure therapy, along with the corresponding total number of patients. Among the 92 patients treated with silicon, there was an average gain of 4.11 points in pain score, 1.61 points in JPG reducibility score, 4.88 points in FAB score, 1.60 points in scar assessment VSS, and 0.78 points in itching score. For the 9 patients treated with soft putty, the average gains were 3.56 points in pain score, 1.67 points in JPG reducibility score, 7.00 points in FAB score, 0.78 points in scar assessment VSS, and 1.89 points in itching score. Among the 45 patients treated with polyethylene foam plastizot, there was an average gain of 4.16 points in pain score, 1.40 points in JPG reducibility score, 5.34 points in FAB score, 1.47 points in scar assessment VSS, and 0.93 points in itching score.

**Table 3:** Outcome score differences associated with different insert materials used.

|  | Total (n) | Average Point Gained in pain score (initial score - final score) | Average Point Gained in Reducibility score (initial score - final score) | Average Point Gained in FAB score (initial score - final score) | Average Point Gained in Scar assessment VSS (initial score - final score) | Average Point Gained in Itching Score (initial score - final score) |
| --- | --- | --- | --- | --- | --- | --- |
| Silicon sheet | 92 | 4.11 | 1.61 | 4.88 | 1.60 | 0.78 |
| Soft putty elastomer effects | 9 | 3.56 | 1.67 | 7.00 | 0.78 | 1.89 |
| Polyethylene foam (Plastazote®) | 45 | 4.16 | 1.40 | 5.34 | 1.47 | 0.93 |

## DISCUSSION

We found that our study underscores the impact of comprehensive physiotherapy treatment on improving patient wellness amidst the challenging backdrop of conflict and resource limitations in Gaza. By focusing on scar hypertrophy management following skin graft surgery, our findings shed light on the efficacy of a tailored rehabilitation program adapted to the unique needs of burn patients, particularly children. One noteworthy aspect of our findings is the association between early intervention, specifically within two weeks post-surgery, and the reliance on tube grip over other pressure therapy tools. This underscores the importance of timely access to rehabilitation services in optimizing patient outcomes, especially in resource-constrained environments and conflict contexts. Furthermore, our observation that silicone sheets were the predominant material used in our program highlights the practical considerations inherent in delivering scar management interventions in conflict settings. Despite the challenges posed by the blockade and limited access to medical supplies, our program has successfully implemented evidence-based practices to address the complex needs of burn patients. Importantly, our study contributes novel insights to the literature on interventions and programs in conflict-affected and resource-limited settings. By demonstrating the feasibility and efficacy of offering comprehensive physiotherapy support in Gaza, we provide a blueprint for similar initiatives grappling with the dual challenges of conflict and constrained resources. Moving forward, it is imperative to leverage our findings to advocate for the expansion and sustainability of such programs in Gaza and beyond. By advocating for increased investment in rehabilitation services within conflict settings, we can ensure equitable access to essential care for those most vulnerable to the enduring repercussions of conflict-related injuries.

The demographic characteristics of our study cohort closely resembled those outlined in a prior descriptive study conducted at the Al Alamy burn center, the primary healthcare facility in the Gaza Strip. This study, published in 2016, offered insights into the epidemiological profile and outcomes among hospitalized burn patients (14). In line with published literature, our study underscores the predominance of pediatric burn injuries, with the upper body emerging as the most commonly affected anatomical region (Ref1). This pattern aligns with existing data attributing a significant portion of pediatric burn injuries to scalds resulting from contact with hot liquids (Ref2). The choice of the thigh as the primary donor site for skin grafts, as observed in our study, reflects established surgical practices aimed at minimizing donor site morbidity and optimizing graft survival (Ref3). This underscores the importance of surgical precision and careful consideration of grafting techniques in achieving favorable outcomes in burn patients.

The timing and initiation of post-surgical physiotherapy are critical determinants of treatment success in burn patients (Ref4). Our study reveals that physiotherapy initiation, occurring approximately two weeks post-surgery on average, aligns with established protocols aimed at ensuring adequate wound healing and minimizing the risk of complications (Ref5). Notably, the protracted duration of rehabilitation, averaging six months in our cohort, underscores the prolonged nature of scar maturation and the importance of sustained therapeutic interventions during this critical period (Ref6). This highlights the need for comprehensive, multidisciplinary rehabilitation programs tailored to the unique needs of burn patients to optimize functional outcomes and quality of life.

Effective scar management is paramount in burn rehabilitation, with pressure therapy and insert materials playing pivotal roles in optimizing scar outcomes (Ref7). Our findings underscore the predominance of tube grip and pressure garments as preferred modalities for pressure therapy, reflecting their effectiveness in minimizing scar contractures and optimizing scar aesthetics (Ref8). However, considerations of cost-effectiveness and resource availability underscore the need for context-specific approaches, particularly in resource-limited settings where alternative materials such as polyethylene foam and soft putty elastomer may offer viable alternatives (Ref9). This highlights the importance of tailoring scar management strategies to individual patient needs and optimizing resource allocation to maximize therapeutic efficacy.

Patient adherence and engagement emerge as central themes in our study, with high rates of treatment compliance and post-treatment follow-up indicative of patient resilience and determination in navigating the complexities of burn care (Ref10). Despite socio-economic adversities and logistical challenges, our findings underscore the importance of patient education, psychosocial support, and community-based interventions in fostering continuity of care and optimizing treatment outcomes (Ref11). This underscores the need for holistic, patient-centered approaches to burn rehabilitation that prioritize patient empowerment and engagement throughout the treatment continuum.

Pressure therapy has emerged as a cornerstone in the management of hypertrophic scars following burn injuries (Ref12). Our study corroborates previous findings regarding the efficacy of pressure garments in scar management, particularly in optimizing scar maturation and minimizing hypertrophic scar development (Ref13). The predominant use of pressure garments in our study population reflects the pragmatic adaptation of standard protocols to resource-constrained settings, highlighting the versatility and effectiveness of pressure therapy in scar management, even in conflict-affected regions like the Gaza Strip (Ref14). Furthermore, our observations regarding recommended pressure levels align with empirical evidence, emphasizing the importance of balancing scar maturation with patient comfort and tolerance (Ref15). However, further research is warranted to refine recommendations regarding pressure duration and intensity, considering the variability in scar response and patient characteristics (Ref16).

Silicone therapy represents another cornerstone in scar management, supported by extensive clinical evidence demonstrating its effectiveness in preventing and treating hypertrophic scars and keloids (Ref17). Our study reaffirms the integration of silicone therapy into comprehensive scar management protocols, particularly in early scar maturation stages (Ref18). The simplicity, non-invasiveness, and favorable side effect profile of silicone products make them invaluable adjuncts to pressure therapy, offering additional benefits in scar pliability, vascularity, and aesthetics (Ref19]. However, ongoing research is needed to elucidate the mechanisms underlying silicone therapy’s efficacy and to optimize treatment protocols, particularly in resource-constrained settings where access to specialized care may be limited (Ref20).

Emerging materials such as thermoplastic polyethylene foam and soft putty elastomer offer promising alternatives to traditional silicone therapy, albeit with limited clinical data (Ref21). Our study underscores the importance of exploring novel materials and technologies to enhance scar management outcomes, particularly in settings where access to specialized resources may be limited (Ref22). Future research endeavors should prioritize rigorous clinical trials to validate the efficacy and safety of these emerging materials, considering factors such as patient comfort, durability, and cost-effectiveness (Ref23). Additionally, interdisciplinary collaboration and knowledge sharing are essential to drive innovation in scar management and improve outcomes for patients worldwide (Ref24).

While our study provides valuable insights into the feasibility and efficacy of pressure therapy and silicone therapy in scar management, several challenges and knowledge gaps remain (Ref25). The socio-political context of the Gaza Strip presents unique challenges to healthcare delivery, including limited access to specialized resources, infrastructure constraints, and political instability (Ref26). Addressing these challenges requires a multifaceted approach, encompassing advocacy, capacity building, and international collaboration to enhance the resilience and sustainability of burn care services in conflict-affected regions (Ref27). Furthermore, future research endeavors should prioritize patient-centered outcomes, community engagement, and implementation science to ensure the effective translation of research findings into clinical practice (Ref28).

The limitations of our study are rooted in its retrospective design and absence of a control group. This inherent design constraint hampers our ability to establish causal relationships between interventions and observed outcomes. Consequently, while our findings illuminate trends in treatment outcomes over time, caution must be taken in attributing these improvements solely to the interventions administered. Moreover, the absence of a control group precludes definitive comparisons between different interventions. Although our study provides valuable insights into the utilization and outcomes associated with pressure therapy methods and insert materials, the lack of a comparative arm limits the robustness of our conclusions. Additionally, the small sample size further underscores the need for cautious interpretation of our findings. With only nine patients receiving soft putty elastomer inserts compared to 92 patients receiving silicone sheets, the uneven distribution of interventions restricts the generalizability of our results. Furthermore, the coexistence of concurrent pressure therapy methods and variability in clinician-driven treatment selection may introduce confounding factors, complicating the interpretation of outcomes. While our study sheds light on the practicalities and outcomes of scar management interventions in a conflict-affected setting, the retrospective design, absence of a control group, and small sample size collectively temper the strength of our conclusions and highlight avenues for future research and improvement. Further research using prospective methods, a prespecified sample size and sufficient power to compare outcomes between treatment groups, would help inform further on which methods, materials, and inserts best support recovery for burn patients in such contexts.

## CONCLUSION

In conclusion, our study underscores the significant role of post-surgical physiotherapy and pressure therapy with insert materials in improving outcomes for patients recovering from burn scars, particularly in conflict-affected regions. Focused on the post-surgical phase following skin graft procedures, our findings illuminate the impact of these interventions on both the physical and psychological aspects of recovery. The integration of physiotherapy and pressure therapy into comprehensive rehabilitation programs represents a promising approach for optimizing scar outcomes and enhancing functional recovery. By facilitating wound healing, minimizing complications, and promoting scar maturation, physiotherapy plays a crucial role in restoring mobility and functional abilities in patients with burn scars. While our study provides valuable insights into the efficacy of these interventions, further research is needed to optimize treatment protocols and refine recommendations for clinical practice. Future studies should focus on elucidating the mechanisms underlying the therapeutic effects of physiotherapy and pressure therapy, as well as evaluating long-term outcomes and cost-effectiveness.

## Declarations

### Ethics approval and consent to participate

This study used retrospectively collected routine programmatic data from Médecins Sans Frontières (MSF) clinical services in Gaza. As the study involved analysis of anonymized routine clinical data, formal ethics committee approval was waived. The study received authorization from the Medical Director of Médecins Sans Frontières Operational Centre Paris and approval from the Gaza Strip Ministry of Health. During their first clinical visit, patients provide written consent allowing the use of their anonymized clinical data for research purposes.

### Availability of data and materials

Data are available upon reasonable request

## Data Availability

The dataset analyzed during the current study contains sensitive clinical patient information from a conflict-affected setting and cannot be made publicly available due to ethical and legal restrictions related to patient confidentiality. De-identified data may be made available upon reasonable request to Médecins Sans Frontières (MSF), subject to approval by the MSF Ethics Review Board and the Gaza Ministry of Health. Requests for access to anonymized data can be directed to the corresponding author.

## Acknowledgements

All the physiotherapists and the medical team in MSF’s outpatient department clinics in Gaza. The managers and data teams who participated in this study. The coordination staff in MSF Operational Centre Paris headquarters for supporting this study. Emma Veitch, freelance medical editor for MSF, provided editorial assistance and her work was funded by MSF-USA.

We also wish to express our sincere thoughts and deep respect for our two colleagues, Abed El Hamid Qaradaya and Hala Al Far, whose dedication and contributions to burn care and rehabilitation in Gaza were invaluable.

## Supplementary materials

**Table S1:**
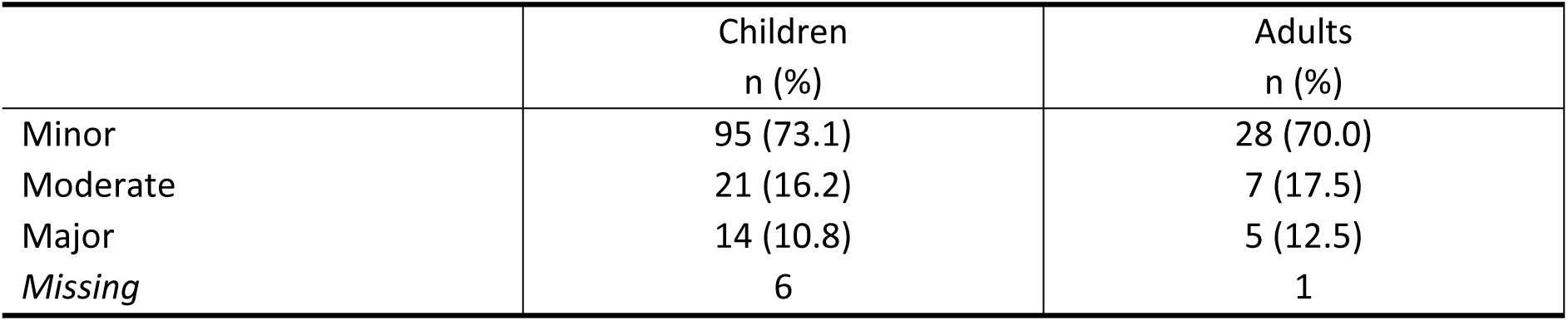
Total surface body area, by age group (child 0-18, Adult 19+).

**Table S2:** Delay between injury, surgery, and admission.

|  | Average | SD |
| --- | --- | --- |
| Average delay between surgery to admission | 13.2 | 75.1 |
| Average delay between admission to discharge | 178.9 | 109.3 |
| Average delay between surgery to discharge | 165.7 | 91.6 |

**Table S3:** Days between admission to physiotherapy and discharge.

|  | Number enrolled | Mean days in care | p25 | p50 | p75 | SD |
| --- | --- | --- | --- | --- | --- | --- |
| **Children** | 136 | 57.09 | 3 | 12 | 46 | 284.20 |
| **Adults** | 41 | 13.68 | 7 | 30 | 53 | 141.73 |

**Table S4:** Scar assessment initial and final (VSS); n(%).

| **Final**<br>**Initial** | **2** | **3** | **4** | **5** | **6** | **7** | **8** | **9** | **10 or +** |
| --- | --- | --- | --- | --- | --- | --- | --- | --- | --- |
| **2** | - | - | - | 1 (50.0) | - | - | - | - | - |
| **3** | - | - | - | - | 1 (50.0) | 1 (50.0) | - | - | - |
| **4** | 1 (11.1) | 2 (22.2) | 5 (55.6) | - | 1 (11.1) | - | - | - | - |
| **5** | - | 2 (11.1) | 8 (44.4) | 5 (27.8) | 3 (16.7) | - | - | - | - |
| **6** | - | 4 (12.5) | 6 (18.8) | 11 (34.4) | 7 (21.9) | - | 4 (12.5) | - | - |
| **7** | 1 (2.8) | 1 (2.8) | 6 (16.7) | 11 (30.6) | 13 (36.1) | 1 (2.8) | 2 (5.6) | - | 1 (2.8) |
| **8** | - | 1 (2.9) | 2 (5.7) | 4 (11.4) | 11 (31.4) | 9 (25.7) | 6 (17.1) | 2 (5.7) | - |
| **9** | - | - | 2 (8.7) | 4 (17.4) | 4 (17.4) | 7 (30.4) | 6 (26.1) | - | - |
| **10 or +** | - | 1 (10.0) | - | - | 1 (10.0) | 1 (10.0) | 5 (50.0) | - | 2 (20.0) |
| **Average initial (SD): 7.1 (1.8)** |  |  |  |  | **Average final (SD): 5.7 (1.7)** |  |  |  |  |

**Table S5:**
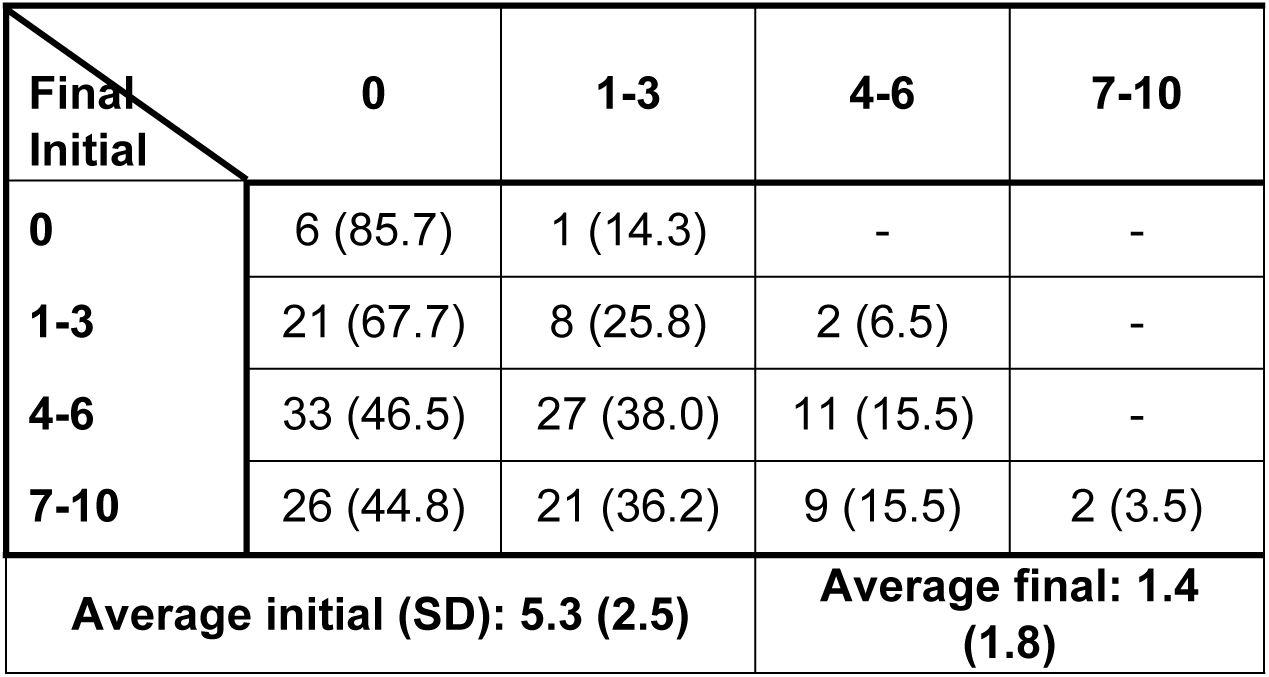
Pain Scores from initial to final (VAS); n (%).

| **Final**<br>**Initial** | **0** | **1-3** | **4-6** | **7-10** |
| --- | --- | --- | --- | --- |
| **0** | 6 (85.7) | 1 (14.3) | - | - |
| **1-3** | 21 (67.7) | 8 (25.8) | 2 (6.5) | - |
| **4-6** | 33 (46.5) | 27 (38.0) | 11 (15.5) | - |
| **7-10** | 26 (44.8) | 21 (36.2) | 9 (15.5) | 2 (3.5) |
| **Average initial (SD): 5.3 (2.5)** |  |  | **Average final: 1.4 (1.8)** |  |

**Table S6:** Itching Scores, initial and final; n (%).

| Final<br>Initial | 0 | 1 | 2 | 3 | 4 | 5 | 6 | 7 | 8 | 9 | 10 |
| --- | --- | --- | --- | --- | --- | --- | --- | --- | --- | --- | --- |
| 0 | 19<br>(47.5) | 2 (5.0) | 5 (12.5) | 6 (15.0) | 3 (7.5) | 3 (7.5) | 1 (2.5) | 1 (2.5) | - | - | - |
| 1 | 2 (50.0) | - | - | 1 (25.0) | 1 (25.0) | - | - | - | - | - | - |
| 2 | 4 (40.0) | 2 (20.0) | 2 (20.0) | 1 (10.0) | - | - | 1 (10.0) | - | - | - | - |
| 3 | 4 (16.0) | 3 (12.0) | 9 (36.0) | 3 (12.0) | 5 (20.0) | - | - | - | 1 (4.0) | - | - |
| 4 | 2 (9.1) | 1 (4.6) | 8 (36.4) | 8 (36.4) | - | - | 2 (9.1) | - | 1 (4.6) | - | - |
| 5 | 1 (4.8) | 1 (4.8) | 6 (28.6) | 6 (28.6) | 2 (8.5) | 3 (14.3) | 2 (9.5) | - | - | - | - |
| 6 | 2 (11.1) | - | 4 (22.2) | 5 (27.8) | 2 (16.7) | 1 (8.3) | 1 (5.6) | - | 1 (5.6) | - | - |
| 7 | 1 (8.3) | 2 (16.7) | 1 (8.3) | 2 (16.7) | 1 (8.3) | 2 (16.7) | 1 (8.3) | 1 (8.3) | 1 (8.3) | - | - |
| 8 | 1 (9.1) | - | - | 3 (27.3) | 4 (36.4) | 1 (9.1) | - | 1 (9.1) | 1 (9.1) | - | - |
| 9 | - | - | - | - | - | 1 (33.3) | 1 (33.3) | 1 (33.3) | - | - | - |
| 10 | - | - | - | - | - | - | - | - | - | - | 1 (100) |
| Average Initial: 3.7 (2.7) |  |  |  |  |  | Average Final: 2.7 (2.2) |  |  |  |  |  |

**Table S7:** Contracture Scores initial and final (JPG); n (%)

| **Final**<br>**Initial** | **0** | **1** | **2** | **3** | **4** | **NA** |
| --- | --- | --- | --- | --- | --- | --- |
| **0** | 21<br>(95.5) | - | - | 1 (4.6) | - | - |
| **1** | 16<br>(59.3) | 11<br>(40.7) | - | - | - | - |
| **2** | 10<br>(28.6) | 16<br>(45.7) | 7 (20.0) | 0 | 1 (2.9) | 1 (2.9) |
| **3** | 12<br>(30.0) | 15<br>(37.5) | 11<br>(27.5) | 2 (5.0) | - | - |
| **4** | 15<br>(32.6) | 15<br>(32.6) | 11<br>(23.9) | 3 (6.5) | 1 (2.2) | 1 (2.2) |
| **NA** | - | 1 (50.0) | - | - | - | 1 (50.0) |
| **Average initial: 2.3 (1.4)** |  |  | **Average final: 0.8 (0.9)** |  |  |  |

**Table S8:** Functional Ability Scores initial and final (FAB); n (%).

| **Final**<br>**Initial** | **High** | **Moderate** | **Mild** | **Dependenc**<br>**e** |
| --- | --- | --- | --- | --- |
| **High dependence** | - | - | - | 3 (100.0) |
| **Moderate dependence** | - | 1 (5.9) | - | 16 (94.1) |
| **Mild dependence** | - | - | - | 20 (100.0) |
| **Dependence** | - | - | - | 36 (100.0) |
| **Average initial: 25.6 (7.1)** |  | **Average final: 34.6 (2.3)** |  |  |

**Figure S1:**
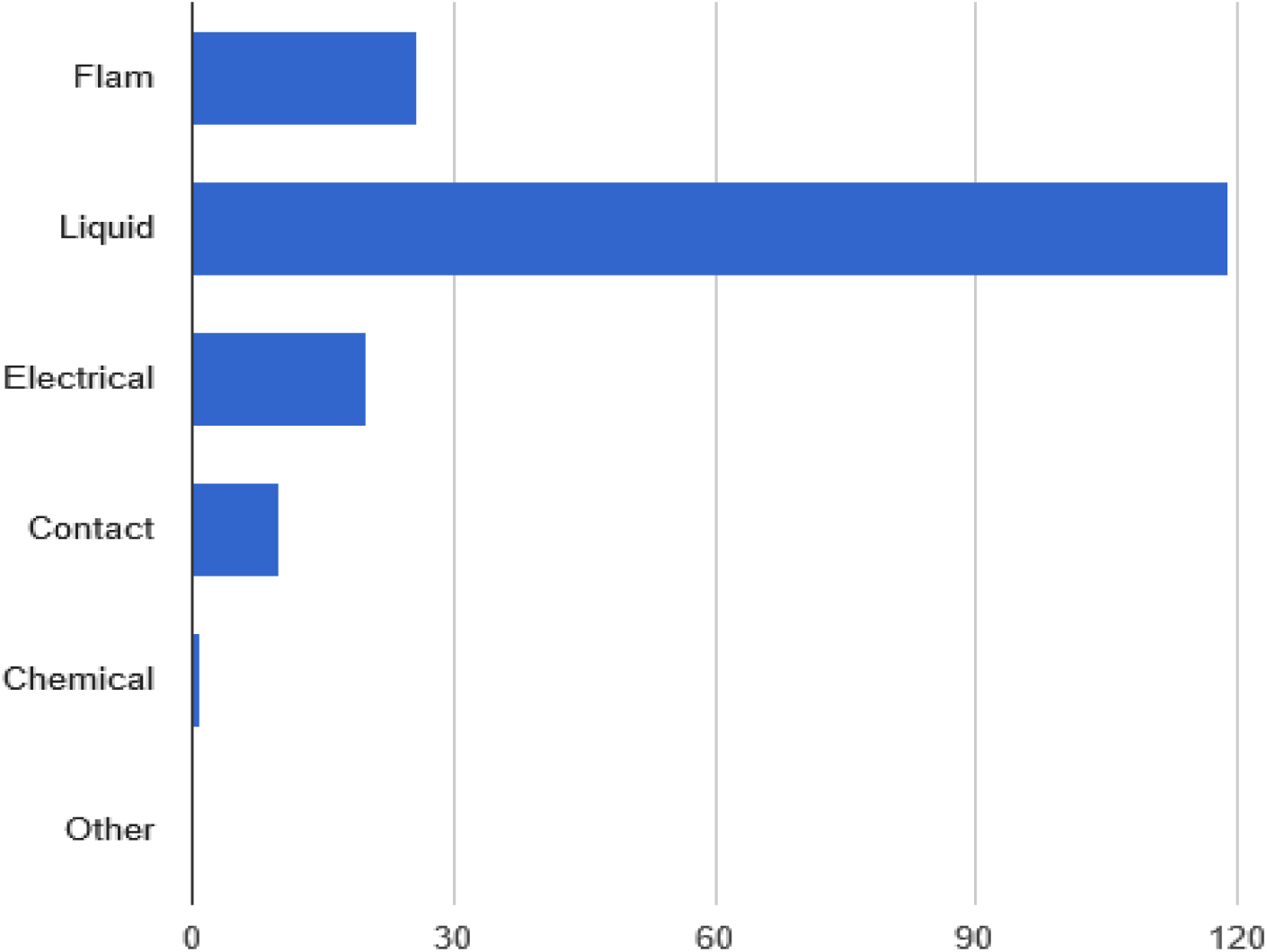
Cause of burn.

**Figure S2:**
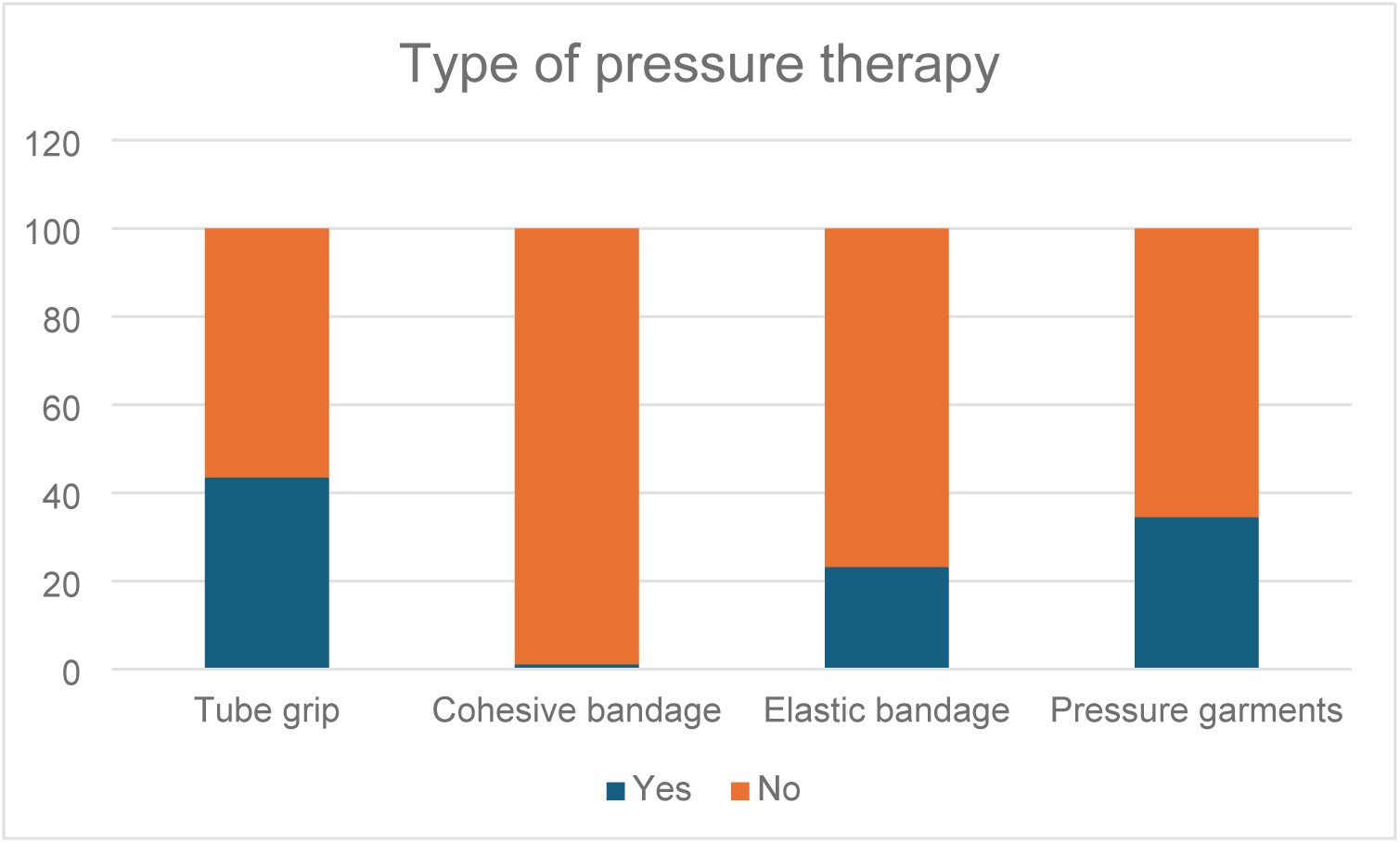
Pressure therapy methods.

